# Healthcare and long-term care for older people with cognitive impairment in the US: A latent transition analysis

**DOI:** 10.1101/2023.10.06.23296615

**Authors:** Bo Hu, Pengyun Wang, Xi Chen

## Abstract

**Background:** Healthcare and long-term care are crucial to the well-being of people with mild cognitive impairment (MCI) or dementia. This study investigates the clustering patterns of healthcare and long-term care use in people with MCI or dementia and the relationships between changes in cognitive functioning and transitions of care.

**Methods:** The study used longitudinal data from three recent waves of the Health and Retirement Study (HRS 2014-2018, N=10,152). MCI and dementia were measured based on the Langa-Weir Classifications. The outcome measures included five types of healthcare and three types of long-term care services. Latent transition analyses were conducted to identify the clustering patterns of care use and map out the transition pathways between different care classes over time. Multilevel regression analyses were conducted to investigate the relationships between changes in cognitive functioning and care transitions.

**Results:** We identified three user groups: medium healthcare and low long-term care (MM-LC, 56%, n=5,653), high healthcare and high long-term care (HM-HC, 37%, n=3,743), and low healthcare and low long-term care (LM-LC, 7%, n=736). The progression of cognitive impairment was associated with a higher probability of transitioning from MM-LC to HM-HC class (β=0.070; p<0.001). An improvement in cognitive functioning was associated with a higher probability of transitioning from HM-HC to MM-LC class (β=0.039, p<0.05).

**Conclusions:** Our findings underscore the importance of integration between healthcare and long-term care. Changes in cognitive functioning are useful indicators for care planning, resource allocation, and care coordination from diverse care providers.

**Key points:**

- Three classes of healthcare and long-term care services were identified in the latent transition analyses.
- The progression of cognitive impairment was associated with transitioning to a class characterized by more use of healthcare and long-term care services,
- An improvement in cognitive functioning was associated with transitioning to a class characterized by less use of care in both sectors.

## Introduction

Cognitive impairment is a broad spectrum of deficits affecting various aspects of cognitive functioning in individuals. Cognitive impairment has various stages, progressing from mild cognitive impairment (MCI) to dementia. The estimated prevalence of MCI among the U.S. population aged 65 years and older is over 20% ^1^. Individuals with MCI have an elevated risk of progression to dementia, the estimated prevalence of which is 10% in adults aged 65 and older. Cognitive impairment is associated with the onset of functional disability and progression of long-term illnesses, leading to complex needs for both health and long-term care. Early identification of cognitive impairment may offer multiple benefits, such as access to treatment, support, and care planning, and delay of institutionalization.^2,3^

Both healthcare and long-term care are crucial to the well-being of people with cognitive impairment. While healthcare services aim to cure a patient’s ill health condition or slow down the progression of the disease, long-term care provided by formal or informal caregivers intends to maintain a person’s functional capability and independence in daily life.^4^ Existing studies have examined the service use of people with MCI or dementia. Overall, a diagnosis of dementia is a major determinant of service use, with increased service use in more severe disease stages, including inpatient admissions,^5^ physician services, home health, skilled nursing facilities,^6,7^ and use of medications.^8^ People with dementia use healthcare services more often than community services.^9–11^ The factors associated with higher levels of service use include impaired ADL,^12^ neuropsychiatric symptoms,^13^ comorbidities,^14^ older age,^15^ having received more education,^16^ not having a spousal caregiver or living alone,^16^ knowledge of available services,^17^ a larger caregiver burden,^18^ and having more care infrastructure.^19^

A substantial evidence gap remains despite the existing research. First, most of the studies focus on single types of service use, while more research is needed to identify how healthcare and long-term care services are combined to meet people’s care needs.^19–21^ Second, studies often examine service use at a particular time point, while less attention is paid to understanding the dynamics of care type transitions.^22^ Finally, previous studies have tended to concentrate on service use in later stages of cognitive impairment, i.e., dementia, while people with MCI already have higher medical costs and receive more informal care compared to people without MCI.^20,23^ It is unclear what types of healthcare and long-term care services are normally being used as cognitive impairment progresses. Yet, such information is crucial if we want to identify and avoid unmet care needs in people with cognitive impairment.

Given the complexity of care needs for people with MCI and dementia, we expect that healthcare and long-term care services are used in combination and cluster into distinct groups. By applying latent transition analysis (LTA), this longitudinal study aims to identify the subgroups of service users and map out their patterns of care transitions over time. Moreover, we hypothesise that the progression of cognitive impairment is strongly associated with care transition. An improved understanding of the relationship between cognitive function and care transition is important for timely access to, and the integration of, healthcare and long-term care for people with MCI or dementia.

## Research methods

### Data and sample

The data used in this paper came from three recent waves of the Health and Retirement Study (HRS), a nationally representative biennial survey of community-dwelling older persons aged 50 years and over in the US (ethical approval: University of Michigan Institutional Review Board).^24^ We focused on those people who participated in the survey and were living with MCI or dementia in 2014, and we followed them until 2018. MCI and dementia were identified by the Langa-Weir Classifications.^25,26^ A cognitive functioning score was derived for each survey participant based on the HRS data. The cutoff points used for classifying individuals into the three categories (i.e., normal, MCI, and dementia) have been reported elsewhere.^25^ The classification algorithms were validated against the prevalence rates reported in the Aging, Demographics, and Memory Study (ADAMS). Though the MCI and dementia classifications are still imperfect, this method accurately identified 78% of HRS respondents’ dementia status (74% for self-respondents and 84% for proxy-respondents).^26^

In 2014, 18,747 persons aged 50 and over participated in the survey, among whom 4,468 persons were identified as living with MCI or dementia in 2014. Of the 4,468 persons, 3,312 and 2,372 persons participated in the survey in 2016 and 2018, respectively. The rest were either deceased or lost to follow-up. Mortality was ascertained through the Respondent’s Exit Interview or Spouse’s Core Interview. If an individual was neither marked as deceased nor had any observable data entries, such an individual was categorized as ‘loss to follow-up’. More details about sample attribution are reported in the Appendix (Figure A1). There were in total 10,152 observations across the three waves (i.e., 4,468+3,312+2,372=10,152). Only a small proportion of observations (<1%) in our sample had missing values.

### Outcome variables

The key outcome variables are the use of healthcare and long-term care services. Following the previous studies ^20^ and subject to data availability in the HRS, we focused on five healthcare services, including doctor visits, prescription medication, hospital inpatient stays, special facilities and services (including adult care center, social worker, outpatient rehabilitation program, physical therapy, and transportation for the elderly or disabled), and outpatient surgery. We examined three types of long-term care, including nursing home services, formal home care, and unpaid care. All healthcare services, nursing home services and formal home care services were ascertained in the HRS for the past two years. The use of unpaid care was restricted to one month before the survey. To focus on long-term nursing home users, we excluded people staying for less than 30 days. Participants were asked whether they had difficulties in performing six activities of daily living (ADLs; dressing, walking, bathing, eating, getting in and out of bed, and using the toilet) and five instrumental activities of daily living (IADLs; cooking, shopping, making phone calls, taking medication, and managing money), and if so, who helped them. Informal care recipients refer to those living at home and receiving help with ADL or IADL difficulties from family members, while formal home care recipients refer to those receiving paid help. Each type of service was coded as a binary variable (0=not using service; 1=service user).

### Key explanatory variables of interest

The cognitive functioning variable had three categories: normal, MCI, and dementia. We tracked the changes in cognitive functioning between two adjacent waves. There were three categories: staying in the same status, progressing in cognitive impairment, and improving in cognitive functioning. The progression of cognitive impairment includes transitioning from normal functioning to MCI or dementia or transitioning from MCI to dementia. An improvement in cognitive functioning refers to transitioning from MCI to normal cognitive functioning.

### Control variables

We controlled for demographic factors, socioeconomic status, and health conditions. Our demographic variables consisted of age, gender, ethnicity, and marital status. The ethnicity variable had three categories: white American, Black/African American, and other ethnicities. The marital status variable was dichotomised: 0=single (never married, separated, divorced, widowed or spouse absent) and 1=married. Regarding socioeconomic status, we focused on years of education (range: 0-17) and annual household income. We controlled for the number of ADL difficulties, IALD difficulties, and chronic diseases.

### Analytic strategy

Our analyses consisted of two stages. Stage one involved identifying the patterns of care utilization and transitions based on latent class analyses (LCA) and latent transition analyses (LTA). The HRS contains rich information on a wide range of care services. LCA/LTA reduces data dimensionality and information complexity, which facilitates resource planning and care coordination in practice.^20,22,27^ We first conducted LCA for each of the three waves to decide the optimal number of classes. We evaluated the log-likelihood, BIC value, entropy, and average posterior probability (APP) of different solutions.^28^ On balance, the three-class model had the most satisfactory model fit and classification quality and thus was considered the optimal solution (Table A1 in Appendix). We then constructed a three-class latent transition model. The model specification can be found in the Appendix. We derived the transition matrices of care utilization from the LTA model. Mortality and loss to follow-up were treated as the absorption states to avoid attrition bias. The LTA model was built using Mplus version 8.4.

Stage two involved converting the transition matrices derived in stage one into dichotomised transition variables (0=staying in the same status and 1=making transitions between two adjacent waves), which were then treated as outcome variables and regressed on the cognitive functioning variables in linear probability models. Random intercepts were added to the regression models to account for intra-individual correlation.^29^ This resulted in a two-level linear probability model. Details about model specifications are reported in the Appendix. While all observations across the three waves of the HRS were used to build the LTA models in stage one, the regression analyses in stage two were based on the transition variables created from two adjacent waves. This means the sample sizes in the two stages of analyses were different.

Attrition and transition are competing events. The attrition sample was not included in the regression models because it contained neither information on the key regressors (i.e., changes in cognitive functioning) nor information on the outcome variable (i.e., transitions in care utilization). Regression analyses were conducted using Stata version 17.

## Results

The characteristics of the pooled sample (HRS 2014-2018) are reported in the second column of Table 1. Sample characteristics in the base year and the two follow-up surveys are reported in Tables A3-A5 in the Appendix. Twenty-nine percent of the pooled sample had dementia, and 54% had MCI (Column 2, Table 1). A detailed breakdown of cognitive functioning by the year of survey is reported in the appendix (Table A2). All of the sample in 2014 had MCI or dementia, among whom 957 and 764 persons reversed to normal cognitive functioning in 2016 and 2018, respectively (Table A2). In total, 17% of the pooled sample were back to normal cognitive functioning (Column 2, Table 1). The average age of the sample was 74.2 years. Fifty-nine percent of the sample were females, and 57% were White Americans. The sample had on average 10.7 years of education, and the mean value of the annual household income was $34,345.

**Table 1.** Sample characteristics, pooled sample of HRS 2014-2018 (N=10,152)

|  | Entire sample | LM-LC | MM-LC | HM-HC |
| --- | --- | --- | --- | --- |
|  | Proportion (%) or mean |  |  |  |
| **Cognitive functioning** |  |  |  |  |
| Normal | 17 | 21 | 21 | 10 |
| MCI | 54 | 61 | 61 | 43 |
| Dementia | 29 | 17 | 18 | 47 |
| **Age** | 74.2 | 67.0 | 72.3 | 78.6 |
| **Gender** |  |  |  |  |
| Male | 41 | 46 | 42 | 37 |
| Female | 59 | 54 | 58 | 63 |
| **Race/ethnicity** |  |  |  |  |
| White American | 57 | 43 | 54 | 64 |
| African American | 32 | 37 | 34 | 27 |
| Other race/ethnicities | 11 | 20 | 12 | 9 |
| **Marital status** |  |  |  |  |
| Single | 60 | 77 | 56 | 63 |
| Married | 40 | 23 | 44 | 37 |
| **Years of education** | 10.7 | 10.2 | 10.6 | 10.9 |
| **Annual household income (\$)** | 34,345 | 28,692 | 35,364 | 33,964 |
| **Number of ADL difficulties (0-6)** | 1.12 | 0.31 | 0.46 | 2.28 |
| **Number IADL difficulties (0-5)** | 1.06 | 0.28 | 0.40 | 2.23 |
| **Number of chronic diseases (0-9)** | 0.25 | 0.14 | 0.22 | 0.33 |
| **n** | 10,152 | 736 | 5,673 | 3,743 |

Notes: LM-LC: low health care and low long-term care; MM-LC: Medium health care and low long-term care; HM-HC: High health care and high long-term care

Table 2 shows the proportion of people who used healthcare and long-term care services broken down by the year of the survey. Around 90% of the sample visited doctors or took prescription medication. Around 40% of the sample reported hospital inpatient stays. One-quarter of the sample used specialist facilities, and one-fifth had had outpatient surgeries. A third of the sample used informal care, whereas 24% used formal home care. More than 10% of the sample had lived in a nursing home for more than 30 days.

**Table 2.** Proportion of people using health and long-term care with 95% Confidence Intervals, HRS 2014-2016 (N=10,152)

|  | 2014 | 2016 | 2018 |
| --- | --- | --- | --- |
| Doctor visits | 90.6 (89.7, 91.4) | 90.1 (89.1, 91.1) | 89.0 (87.7, 90.2) |
| Prescription medication | 92.6 (91.7, 93.3) | 92.3 (91.4, 93.2) | 92.7 (91.6, 93.7) |
| Hospital inpatient stays | 43.0 (41.6, 44.5) | 43.0 (41.4, 44.7) | 39.1 (37.1, 41.0) |
| Specialist facilities | 24.4 (23.2, 25.7) | 25.0 (23.6, 26.5) | 24.0 (22.3, 25.8) |
| Outpatient surgery | 20.1 (18.9, 21.3) | 20.6 (19.3, 22.0) | 18.8 (17.2, 20.4) |
| Informal care | 33.8 (32.4, 35.2) | 32.8 (31.3, 34.4) | 32.0 (30.2, 33.9) |
| Formal care | 24.2 (23.0, 25.5) | 25.1 (23.6, 26.6) | 23.4 (21.8, 25.2) |
| Nursing home | 15.5 (14.4, 16.5) | 15.0 (13.8, 16.2) | 11.8 (10.6, 13.2) |
| **n** | 4,468 | 3,312 | 2,372 |

The results of the three-class LTA model are reported in Figure 1. People in class 1, labelled as the low healthcare and low long-term care class (LM-LC), had a moderate probability of visiting doctors and a low probability of using all other care services. People in class 2 were labelled as the medium healthcare and low long-term care class (MM-LC). This class was characterised by a high probability of visiting doctors and taking medication and a moderate probability of having inpatient stays and outpatient surgeries. People in class 3 had a high probability of using all care services and were labelled as the high healthcare high long-term care class (HM-HC). The MM-LC class was the major class, with 56% of the sample (n=5,673) belonging to this group. While 37% of the sample (n=3,743) belonged to the HM-HC group, only 7% (n=736) were classified into the LM-LC group. The sample characteristics broken down by the three latent classes are reported in columns 3-5 in Table 1.

**Figure 1.**
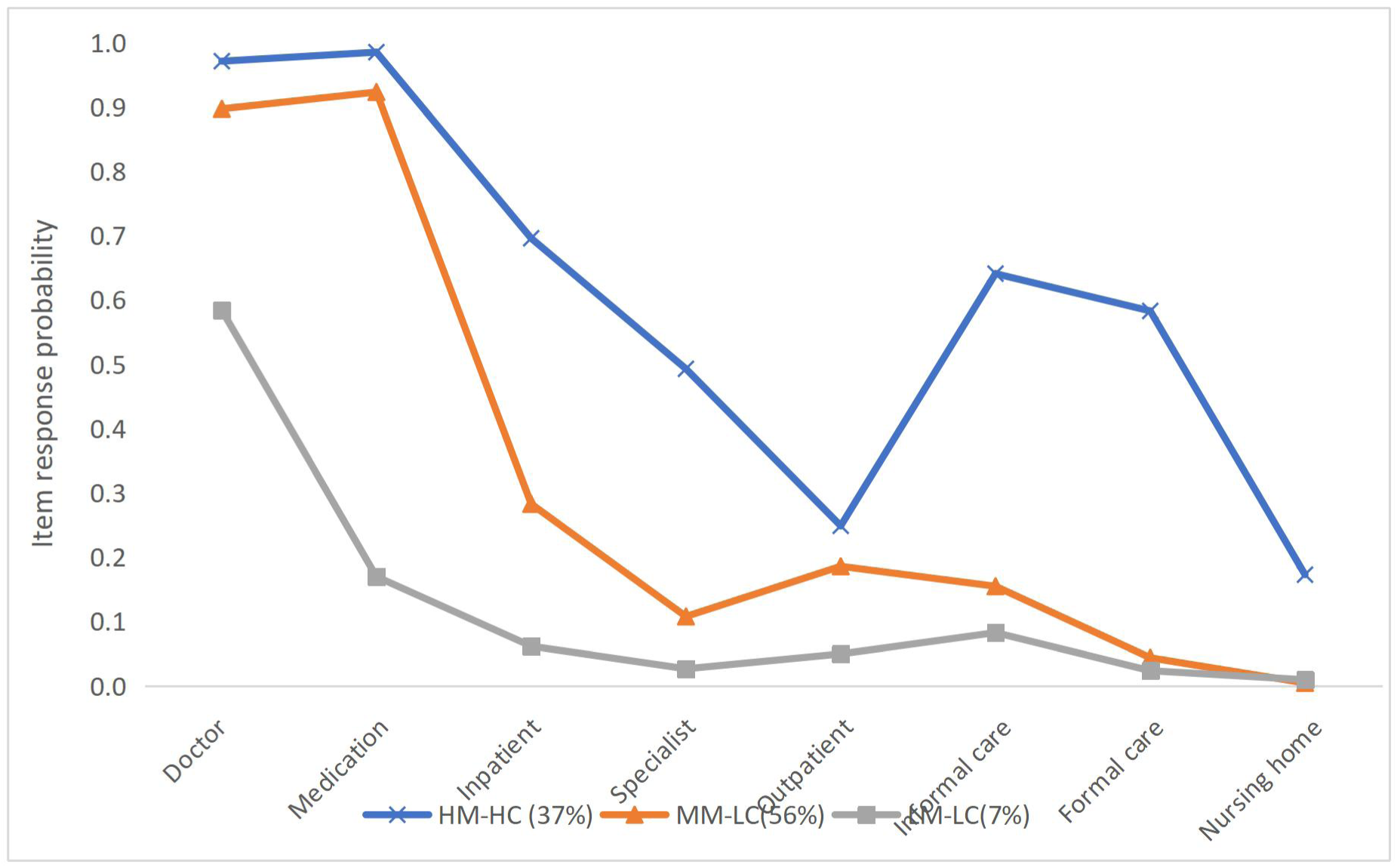
Item response probabilities of the latent transition model, HRS 2014-2018 (N=10,152) Notes: LM-LC: low health care and low long-term care; MM-LC: Medium health care and low long-term care; HM-HC: High health care and high long-term care

Table 3 shows the probabilities of transitions between the different classes. The raw numbers underlying the transition probabilities are reported in Table A6 in the Appendix. Among the 334 people in the LM-LC class in 2014, 15% moved to the MM-LC class and 3.0% moved to the HM-HC class in 2016. There were 240 people in the LM-LC class in 2016, among whom 3.3% moved to the MM-LC class and 3.8% moved to the HM-HC class in 2018. Among the 2,428 people in the MM-LC class in 2014, 9.8% moved to the HM-HC class in 2016. Among the 1,839 people in the MM-LC class in 2016, 6.0% moved to the HM-HC class in 2018. Among the 1,706 people in the HM-HC class in 2014, 3.8% moved to the MM-LC class in 2016, and among the 1,233 people in the HM-HC class in 2016, 4.8% moved to the MM-LC class in 2018.

**Table 3.** Transition matrix of healthcare and long-term care utilization, HRS 2014-2018 (N=10,152)

| 2016 |  |  |  |  |  |  |
| --- | --- | --- | --- | --- | --- | --- |
| 2014 | LM-LC | MM-LC | HM-HC | Mortality | Loss to follow up | Total number |
| LM-LC | 67.4% | 15.0% | 3.0% | 3.9% | 10.8% | 334 |
| MM-LC | 0.3% | 70.6% | 9.8% | 10.5% | 8.7% | 2,428 |
| HM-HC | 0.4% | 4.4% | 57.7% | 30.3% | 7.2% | 1,706 |
| 2018 |  |  |  |  |  |  |
| 2016 | LM-LC | MM-LC | HM-HC | Mortality | Loss to follow up | Total number |
| LM-LC | 62.5% | 3.3% | 3.8% | 4.2% | 26.3% | 240 |
| MM-LC | 0.1% | 70.6% | 6.0% | 8.0% | 15.2% | 1,839 |
| HM-HC | 0.3% | 4.8% | 54.4% | 24.6% | 15.9% | 1,233 |

Notes: LM-LC: low healthcare and low long-term care; MM-LC: Medium healthcare and low long-term care; HM-HC: High healthcare and high long-term care

Three transition pathways were most common in the transition matrices: (1) from the LM-LC class to either the MM-LC or HM-HC class vs. staying in the LM-LC class; (2) from the MM-LC class to the HM-HC class vs. staying in the MM-LC class; and (3) from the HM-HC class to the MM-LC class vs. staying in the HM-HC class. These three pathways were converted into three binary outcome variables, which were then respectively regressed on the transitions of cognitive functioning and control variables in three regression models 1-3 (Table 4). Since the proportions of people in other transition pathways are negligible (<0.5%), they were not further investigated.

**Table 4.**
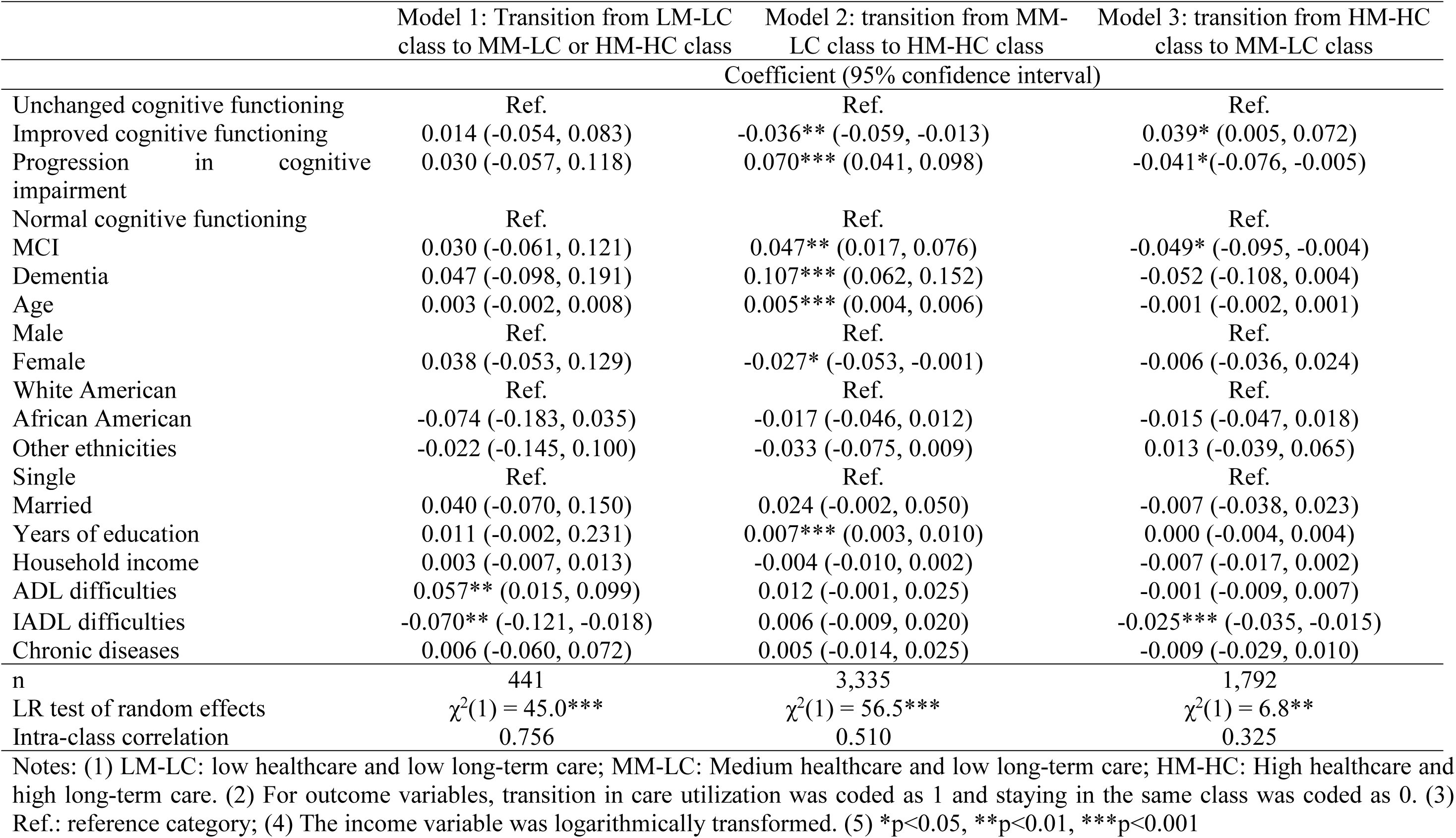
Association between changes in cognitive functioning and transitions in the utilization of health and long-term care, two-level linear probability models.

|  | Model 1: Transition from LM-LC class to MM-LC or HM-HC class | Model 2: transition from MM-LC class to HM-HC class | Model 3: transition from HM-HC class to MM-LC class |
| --- | --- | --- | --- |
|  | Coefficient (95% confidence interval) |  |  |
| Unchanged cognitive functioning | Ref. | Ref. | Ref. |
| Improved cognitive functioning | 0.014 (-0.054, 0.083) | -0.036** (-0.059, -0.013) | 0.039* (0.005, 0.072) |
| Progression in cognitive impairment | 0.030 (-0.057, 0.118) | 0.070*** (0.041, 0.098) | -0.041*(-0.076, -0.005) |
| Normal cognitive functioning | Ref. | Ref. | Ref. |
| MCI | 0.030 (-0.061, 0.121) | 0.047** (0.017, 0.076) | -0.049* (-0.095, -0.004) |
| Dementia | 0.047 (-0.098, 0.191) | 0.107*** (0.062, 0.152) | -0.052 (-0.108, 0.004) |
| Age | 0.003 (-0.002, 0.008) | 0.005*** (0.004, 0.006) | -0.001 (-0.002, 0.001) |
| Male | Ref. | Ref. | Ref. |
| Female | 0.038 (-0.053, 0.129) | -0.027* (-0.053, -0.001) | -0.006 (-0.036, 0.024) |
| White American | Ref. | Ref. | Ref. |
| African American | -0.074 (-0.183, 0.035) | -0.017 (-0.046, 0.012) | -0.015 (-0.047, 0.018) |
| Other ethnicities | -0.022 (-0.145, 0.100) | -0.033 (-0.075, 0.009) | 0.013 (-0.039, 0.065) |
| Single | Ref. | Ref. | Ref. |
| Married | 0.040 (-0.070, 0.150) | 0.024 (-0.002, 0.050) | -0.007 (-0.038, 0.023) |
| Years of education | 0.011 (-0.002, 0.231) | 0.007*** (0.003, 0.010) | 0.000 (-0.004, 0.004) |
| Household income | 0.003 (-0.007, 0.013) | -0.004 (-0.010, 0.002) | -0.007 (-0.017, 0.002) |
| ADL difficulties | 0.057** (0.015, 0.099) | 0.012 (-0.001, 0.025) | -0.001 (-0.009, 0.007) |
| IADL difficulties | -0.070** (-0.121, -0.018) | 0.006 (-0.009, 0.020) | -0.025*** (-0.035, -0.015) |
| Chronic diseases | 0.006 (-0.060, 0.072) | 0.005 (-0.014, 0.025) | -0.009 (-0.029, 0.010) |
| n | 441 | 3,335 | 1,792 |
| LR test of random effects | $\chi^2(1) = 45.0***$ | $\chi^2(1) = 56.5***$ | $\chi^2(1) = 6.8**$ |
| Intra-class correlation | 0.756 | 0.510 | 0.325 |

Notes: (1) LM-LC: low healthcare and low long-term care; MM-LC: Medium healthcare and low long-term care; HM-HC: High healthcare and high long-term care. (2) For outcome variables, transition in care utilization was coded as 1 and staying in the same class was coded as 0. (3) Ref.: reference category; (4) The income variable was logarithmically transformed. (5) \*p<0.05, \*\*p<0.01, \*\*\*p<0.001

Model 2 in Table 4 shows that an improvement in cognitive functioning was associated with a lower probability (β=-0.036, p<0.01) of transitioning from the MM-LC to HM-HC class than staying in the MM-LC class. Model 3 shows that cognitive functioning improvement was associated with a higher probability of transitioning from the HM-HC to MM-LC class than staying in the HM-HC class (β=0.039, p<0.05). People experiencing a progression in cognitive impairment had a higher probability to transition from the MM-LC to HM-HC class than to stay in the MM-LC class in model 2 (β=0.070, p<0.001); and a lower probability to transition from the HM-HC to MM-LC class than to stay in the HM-HC in model 3 (β=-0.041, p<0.05). There is no strong evidence that changes in cognitive functioning were strongly associated with transitions from the LM-LC to other classes (Model 1).

The initial status of cognitive functioning matters. People with MCI or dementia in the previous wave were more likely to transition from the MM-LC class to the HM-HC class by the next wave. In addition, the probability of transitions from the MM-LC class to the HM-HC class was lower among females and higher among older adults and people receiving more years of education. Both the likelihood ratio test of the random intercept and the coefficient of intra-class correlation suggest that individual-level transitions were strongly correlated over time and thus should be accounted for in a multilevel model.

## Discussion

Drawing on longitudinal data from a nationally representative survey, this study investigated the trajectories of healthcare and long-term care utilization in people with MCI or dementia in the US. The latent transition analyses show that people with cognitive impairment did not choose care forms randomly but used specific combinations of services to meet their complex care needs, which is consistent with findings from other countries.^20,27^

Although more than half of the sample stayed in the same care utilization group, which indicated a degree of stability over time, a noticeable proportion transitioned to other groups. It was common that people transitioned from the LM-LC group to the MM-LC group or from the MM-LC group to the HM-HC group, which represented more engagement with healthcare and long-term care services. Meanwhile, a decline in the utilization of care services was also observed with around 4.5% of the sample transitioning from the HM-HC group to the MM-LC group.

We found that the progression of cognitive impairment was often accompanied by a heightened utilization of healthcare and long-term care, whereas an improvement in cognitive functioning went in tandem with a decrease in care utilization. Care needs are the most immediate driver of care use.^12–14^ While many existing studies have investigated the need for healthcare and long-term care separately, our study shows that cognitive impairment is a strong driver of care utilization in both sectors. More importantly, the current level of and changes in cognitive functioning not only indicate the existing need for care services but also provide valuable information about how care utilization might change in the future.

Our research findings have important policy implications. First, the combined use of healthcare and long-term care among people with MCI or dementia brings to the fore the importance of care integration and coordination. Meaningful collaboration, information sharing facilitated by communication technology, joint training, and the designation of care coordinators between the two sectors are fundamental to healthcare and long-term care integration.^21,30^ Integrated care is expected to play an increasingly significant role considering that the number of people with MCI or dementia will continue to rise with population ageing.^1^

Second, practitioners and policymakers may want to closely monitor individuals’ changes in cognitive functioning over time and make plans regarding care arrangements accordingly. Early planning allows for sufficient time for resource allocation, which promotes the timely delivery of personalised care and decreases the risks of unmet care needs.

Finally, our analyses showed that continued increases in care demand are not inevitable outcomes in people with cognitive impairment. Healthy ageing policies, especially those promoting brain health and improving the detection or prevention of MCI, hold great potential in addressing the economic challenges posed to the health and long-term care systems across the world.

The limitations of the study should be acknowledged. First, our study reported a strong association between changes in cognitive functioning and transitions in care utilization, while disentangling the direction of causality will be challenging. Second, the analyses focused on whether people used individual services, but did not touch upon the frequency and intensity of care use. Finally, care utilization was based on self-reported and recalled information, which can be subject to recall bias.

## Conclusion

People with MCI or dementia form distinct groups in the utilization of healthcare and long-term care. Changes in cognitive functioning are among the most important predictors of care transitions and thus can be useful indicators for policymakers in terms of facilitating care integration, optimizing resource allocation, and coordinating care supply from diverse providers.

## Data Availability

All data produced in the present study are available upon reasonable request to the authors
All data produced in the present work are contained in the manuscript
All data produced are available online at Health and Retirement Study (HRS)

https://www.rand.org/well-being/social-and-behavioral-policy/centers/aging/dataprod.html

https://hrsdata.isr.umich.edu/

## Appendix

### Specification of the statistical models

The latent transition model in the study was specified as follows:

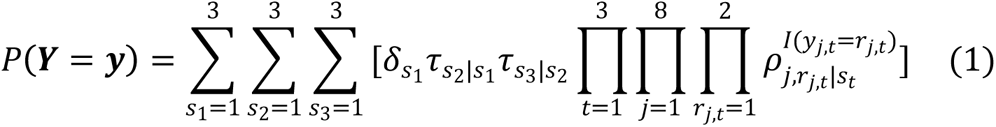

Where *P*(*Y* = *y*) denotes the joint probabilities of a vector of responses for the eight types of care services across three time points, 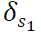 denotes the probabilities of membership in each latent class at time 1, and 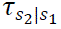 denotes the probabilities of memberships in each latent class at time 2 conditional upon belonging to a particular latent class at time 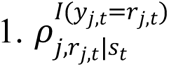 denotes the time-specific probability of using a particular type of service conditional upon belonging to a latent class (i.e., item response probability). Item response probabilities were assumed to be equal across time (namely, 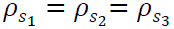) for model parsimony.^28^ We derived the transition matrices from the LTA model. Death and loss-to-follow-up were included in the LTA models and treated as absorbing states in the transition matrices.

We specified the two-level linear probability models as follows:

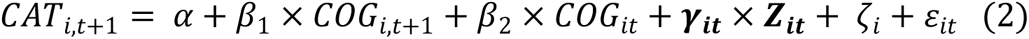

Where 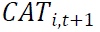 is a particular type of care transition for individual *i* from time point t to time point *t* + 1 (*t* = 1,2), *COG_i,t_*_+1_ denotes the transitioning in cognitive functioning from *t* to t+1, *COG_it_* denotes the status of cognitive functioning at time point *t*, and ***Z****_it_* is a vector of control variables. *β*_1_, *β*_2_, and ***y****_i_* are their respective coefficients. ζ*_i_* denotes the level-two individual-specific error (i.e., random intercept) and ε*_it_* is the level-one error term.

**Table A1.** Model fit and classification quality of exploratory latent class models.

| **2014** |  |  |  |  |
| --- | --- | --- | --- | --- |
|  | Two-group<br>model | Three-group<br>model | Four-group<br>model | Five-group<br>model |
| **Log-likelihood** | -15722 | -15557 | -15510 | -15481 |
| **BIC** | 31587 | 31333 | 31315 | 31333 |
| **Entropy** | 0.633 | 0.610 | 0.573 | 0.561 |
| **APP** |  |  |  |  |
| Group 1 | 0.894 | 0.909 | 0.753 | 0.561 |
| Group 2 | 0.906 | 0.788 | 0.844 | 0.698 |
| Group 3 |  | 0.858 | 0.772 | 0.800 |
| Group 4 |  |  | 0.695 | 0.643 |
| Group 5 |  |  |  | 0.701 |
| **2016** |  |  |  |  |
|  | Two-group<br>model | Three-group<br>model | Four-group<br>model | Five-group<br>model |
| **Log-likelihood** | -11547 | -11450 | -11414 | -11399 |
| **BIC** | 23232 | 23111 | 23113 | 23154 |
| **Entropy** | 0.611 | 0.603 | 0.509 | 0.499 |
| **APP** |  |  |  |  |
| Group 1 | 0.901 | 0.890 | 0.844 | 0.640 |
| Group 2 | 0.880 | 0.805 | 0.773 | 0.617 |
| Group 3 |  | 0.802 | 0.846 | 0.750 |
| Group 4 |  |  | 0.618 | 0.903 |
| Group 5 |  |  |  | 0.623 |
| **2018** |  |  |  |  |
|  | Two-group<br>model | Three-group<br>model | Four-group<br>model | Five-group<br>model |
| **Log-likelihood** | -8252 | -8199 | -8171 | -8153 |
| **BIC** | 16636 | 16600 | 16614 | 16648 |
| **Entropy** | 0.590 | 0.593 | 0.671 | 0.646 |
| **APP** |  |  |  |  |
| Group 1 | 0.871 | 0.818 | 0.807 | 0.702 |
| Group 2 | 0.890 | 0.827 | 0.833 | 0.695 |
| Group 3 |  | 0.741 | 0.829 | 0.743 |
| Group 4 |  |  | 0.801 | 0.647 |
| Group 5 |  |  |  | 0.834 |

Note: APP: average posterior probability

**Table A2.** Number of people in the three cognitive groups in the base line and follow-up surveys.

|  | **2014** | **2016** | **2018** |
| --- | --- | --- | --- |
| **Normal** | - | 957 | 764 |
| **MCI** | 3,156 | 1,404 | 947 |
| **Dementia** | 1,312 | 951 | 661 |
| **Mortality** | - | 784 | 1,245 |
| **Loss to follow up** | - | 372 | 851 |
| **n** | 4,468 | 4,468 | 4,468 |

**Table A3.**
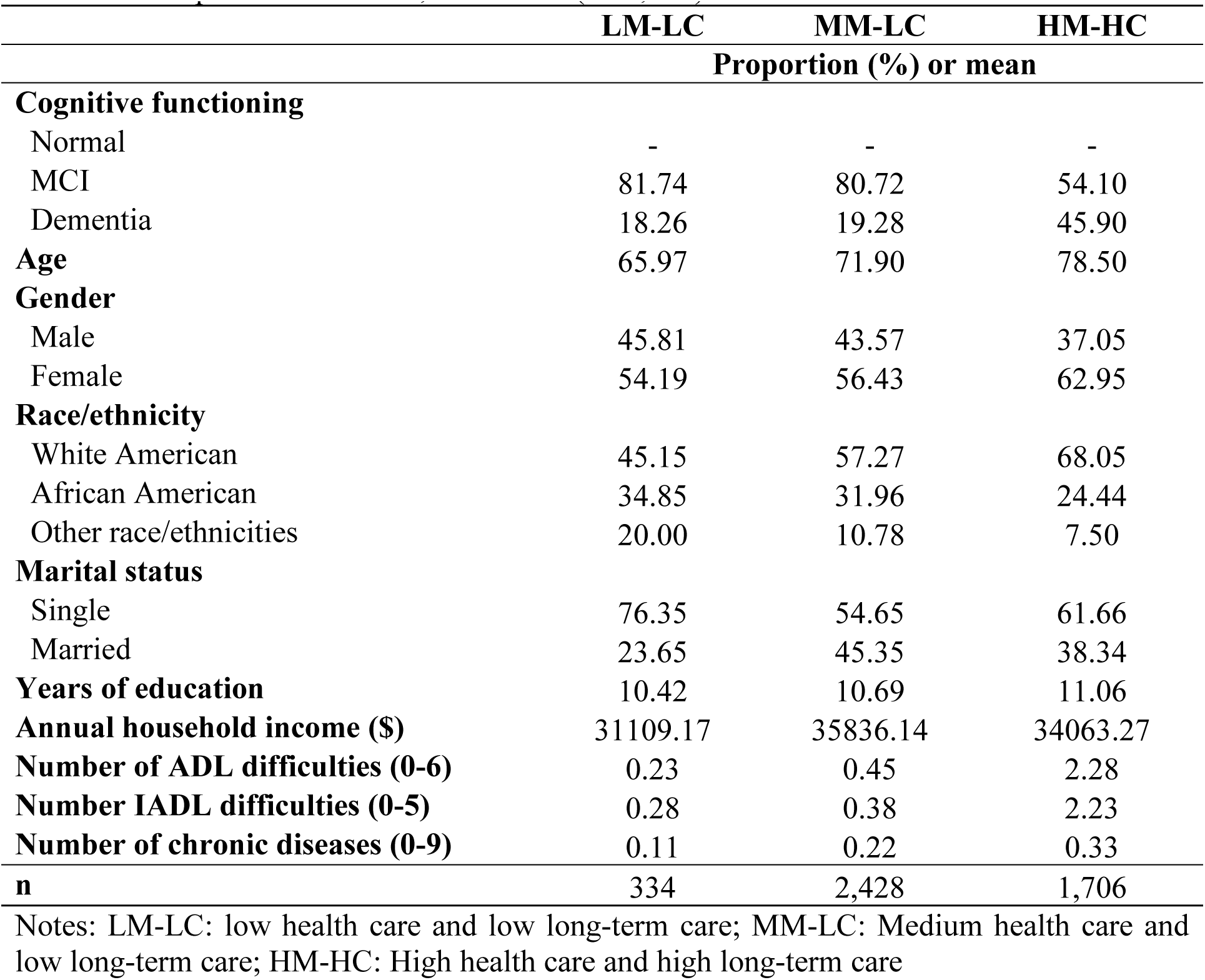
Sample characteristics, HRS 2014 (N=4,468)

|  | LM-LC | MM-LC | HM-HC |
| --- | --- | --- | --- |
|  | Proportion (%) or mean |  |  |
| **Cognitive functioning** |  |  |  |
| Normal | - | - | - |
| MCI | 81.74 | 80.72 | 54.10 |
| Dementia | 18.26 | 19.28 | 45.90 |
| **Age** | 65.97 | 71.90 | 78.50 |
| **Gender** |  |  |  |
| Male | 45.81 | 43.57 | 37.05 |
| Female | 54.19 | 56.43 | 62.95 |
| **Race/ethnicity** |  |  |  |
| White American | 45.15 | 57.27 | 68.05 |
| African American | 34.85 | 31.96 | 24.44 |
| Other race/ethnicities | 20.00 | 10.78 | 7.50 |
| **Marital status** |  |  |  |
| Single | 76.35 | 54.65 | 61.66 |
| Married | 23.65 | 45.35 | 38.34 |
| **Years of education** | 10.42 | 10.69 | 11.06 |
| **Annual household income (\$)** | 31109.17 | 35836.14 | 34063.27 |
| **Number of ADL difficulties (0-6)** | 0.23 | 0.45 | 2.28 |
| **Number IADL difficulties (0-5)** | 0.28 | 0.38 | 2.23 |
| **Number of chronic diseases (0-9)** | 0.11 | 0.22 | 0.33 |
| **n** | 334 | 2,428 | 1,706 |

Notes: LM-LC: low health care and low long-term care; MM-LC: Medium health care and low long-term care; HM-HC: High health care and high long-term care

**Table A4.** Sample characteristics, HRS 2016 (N=3,312)

|  | LM-LC | MM-LC | HM-HC |
| --- | --- | --- | --- |
|  | Proportion (%) or mean |  |  |
| **Cognitive functioning** |  |  |  |
| Normal | 37.92 | 35.62 | 17.11 |
| MCI | 46.67 | 47.20 | 34.39 |
| Dementia | 15.42 | 17.18 | 48.50 |
| **Age** | 66.96 | 72.42 | 78.68 |
| **Gender** |  |  |  |
| Male | 47.50 | 41.49 | 37.79 |
| Female | 52.50 | 58.51 | 62.21 |
| **Race/ethnicity** |  |  |  |
| White American | 42.37 | 53.95 | 63.96 |
| African American | 37.29 | 34.01 | 27.84 |
| Other race/ethnicities | 20.34 | 12.04 | 8.20 |
| **Marital status** |  |  |  |
| Single | 77.08 | 55.01 | 63.99 |
| Married | 22.92 | 44.99 | 36.01 |
| **Years of education** | 10.13 | 10.52 | 10.88 |
| **Annual household income (\$)** | 26663.16 | 35467.70 | 32714.61 |
| **Number of ADL difficulties (0-6)** | 0.36 | 0.46 | 2.30 |
| **Number IADL difficulties (0-5)** | 0.28 | 0.42 | 2.24 |
| **Number of chronic diseases (0-9)** | 0.16 | 0.24 | 0.37 |
| **n** | 240 | 1,839 | 1,233 |

Notes: LM-LC: low health care and low long-term care; MM-LC: Medium health care and low long-term care; HM-HC: High health care and high long-term care

**Table A5.** Sample characteristics, HRS 2018 (N=2,372)

|  | LM-LC | MM-LC | HM-HC |
| --- | --- | --- | --- |
|  | Proportion (%) or mean |  |  |
| **Cognitive functioning** |  |  |  |
| Normal | 41.36 | 38.12 | 20.02 |
| MCI | 41.36 | 44.03 | 32.46 |
| Dementia | 17.28 | 17.85 | 47.51 |
| **Age** | 69.19 | 72.91 | 78.61 |
| **Gender** |  |  |  |
| Male | 45.68 | 40.11 | 36.19 |
| Female | 54.32 | 59.89 | 63.81 |
| **Race/ethnicity** |  |  |  |
| White American | 40.88 | 49.50 | 58.83 |
| African American | 38.99 | 37.38 | 32.09 |
| Other race/ethnicities | 20.13 | 13.12 | 9.08 |
| **Marital status** |  |  |  |
| Single | 77.78 | 57.75 | 63.31 |
| Married | 22.22 | 42.25 | 36.69 |
| **Years of education** | 9.93 | 10.42 | 10.84 |
| **Annual household income (\$)** | 26712.78 | 34146.33 | 35667.97 |
| **Number of ADL difficulties (0-6)** | 0.39 | 0.48 | 2.25 |
| **Number IADL difficulties (0-5)** | 0.30 | 0.42 | 2.19 |
| **Number of chronic diseases (0-9)** | 0.17 | 0.20 | 0.27 |
| **n** | 162 | 1,406 | 804 |

Notes: LM-LC: low health care and low long-term care; MM-LC: Medium health care and low long-term care; HM-HC: High health care and high long-term care

**Table A6.** Number of people making transitions in the utilization of healthcare and long-term care, HRS 2014-2018.

| 2016 |  |  |  |  |  |  |
| --- | --- | --- | --- | --- | --- | --- |
| 2014 | LM-LC | MM-LC | HM-HC | Mortality | Loss to follow up | Total |
| LM-LC | 225 | 50 | 10 | 13 | 36 | 334 |
| MM-LC | 8 | 1,714 | 238 | 256 | 212 | 2,428 |
| HM-HC | 7 | 75 | 985 | 517 | 122 | 1,706 |
| 2018 |  |  |  |  |  |  |
| 2016 | LM-LC | MM-LC | HM-HC | Mortality | Loss to follow up | Total |
| LM-LC | 150 | 8 | 9 | 10 | 63 | 240 |
| MM-LC | 1 | 1,299 | 111 | 148 | 280 | 1,839 |
| HM-HC | 4 | 59 | 671 | 303 | 196 | 1,232 |

**Figure A1.**
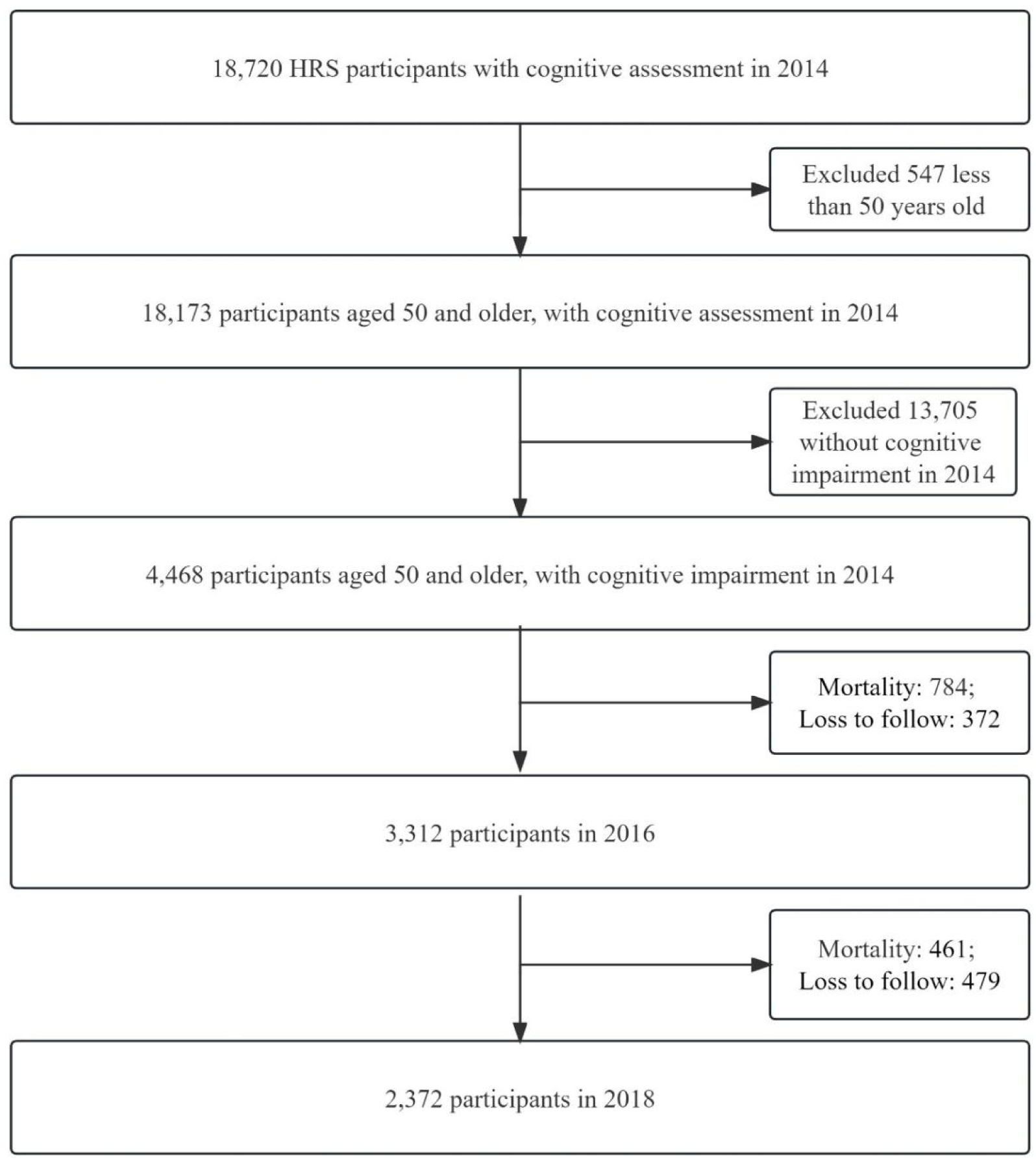
Flow Chart: Analytical sample using three waves of HRS (2014, 2016, 2018)

## Reference

1. Manly JJ, Jones RN, Langa KM, et al. Estimating the prevalence of dementia and mild cognitive impairment in the US: The 2016 Health and Retirement Study Harmonized Cognitive Assessment Protocol Project. 2022;79(12):1242–1249. doi: 10.1001/jamaneurol.2022.3543

2. Giezendanner S, Monsch AU, Kressig RW, et al. General practitioners’ attitudes towards early diagnosis of dementia: a cross-sectional survey. BMC Fam Pract. 2019;20(1):65. doi: 10.1186/s12875-019-0956-1

3. Patnode CD, Perdue LA, Rossom RC, et al. Screening for Cognitive Impairment in Older Adults: Updated Evidence Report and Systematic Review for the US Preventive Services Task Force. JAMA. 2020;323(8):764–785. doi: 10.1001/jama.2019.22258

4. Verbrugge LM, Jette AM. The disablement process. Soc Sci Med. 1994;38(1):1-14. doi: 10.1016/0277-9536(94)90294-1

5. Zhu CW, Cosentino S, Ornstein K, et al. Medicare Utilization and Expenditures Around Incident Dementia in a Multiethnic Cohort. J Gerontol A Biol Sci Med Sci. 2015;70(11):1448–1453. doi: 10.1093/gerona/glv124

6. Hoffman GJ, Maust DT, Harris M, Ha J, Davis MA. Medicare spending associated with a dementia diagnosis among older adults. J Am Geriatr Soc. 2022;70(9):2592–2601. doi: 10.1111/jgs.17835

7. Lin PJ, Zhong Y, Fillit HM, Chen E, Neumann PJ. Medicare Expenditures of Individuals with Alzheimer’s Disease and Related Dementias or Mild Cognitive Impairment Before and After Diagnosis. J Am Geriatr Soc. 2016;64(8):1549–1557. doi: 10.1111/jgs.14227

8. Rabinovici GD, Gatsonis C, Apgar C, et al. Association of Amyloid Positron Emission Tomography With Subsequent Change in Clinical Management Among Medicare Beneficiaries With Mild Cognitive Impairment or Dementia. JAMA. 2019;321(13):1286–1294. doi: 10.1001/jama.2019.2000

9. Chung SD, Liu SP, Sheu JJ, Lin CC, Lin HC, Chen CH. Increased healthcare service utilizations for patients with dementia: a population-based study. PLoS One. 2014;9(8):e105789. doi: 10.1371/journal.pone.0105789

10. Chen L, Reed C, Happich M, Nyhuis A, Lenox-Smith A. Health care resource utilisation in primary care prior to and after a diagnosis of Alzheimer’s disease: a retrospective, matched case-control study in the United Kingdom. BMC Geriatr. 2014;14:76. doi: 10.1186/1471-2318-14-76

11. Weber SR, Pirraglia PA, Kunik ME. Use of services by community-dwelling patients with dementia: a systematic review. Am J Alzheimers Dis Other Demen. 2011;26(3):195–204. doi: 10.1177/1533317510392564

12. Bergvall N, Brinck P, Eek D, et al. Relative importance of patient disease indicators on informal care and caregiver burden in Alzheimer’s disease. Int Psychogeriatr. 2011;23(1):73–85. doi: 10.1017/S1041610210000785

13. Jonsson L, Eriksdotter Jonhagen M, Kilander L, et al. Determinants of costs of care for patients with Alzheimer’s disease. Int J Geriatr Psychiatry. 2006;21(5):449–459. doi: 10.1002/gps.1489

14. Scalmana S, Di Napoli A, Franco F, et al. Use of health and social care services in a cohort of Italian dementia patients. Funct Neurol. 2013;28(4):265–273. doi:

15. Boersma F, Eefsting JA, van den Brink W, van Tilburg W. Care services for dementia patients: predictors for service utilization. Int J Geriatr Psychiatry. 1997;12(11):1119–1126. doi: 10.1002/(sici)1099-1166(199711)12:11<1119::aid-gps702>3.0.co;2-h

16. Toseland RW, McCallion P, Gerber T, Banks S. Predictors of health and human services use by persons with dementia and their family caregivers. Soc Sci Med. 2002;55(7):1255–1266. doi: 10.1016/s0277-9536(01)00240-4

17. Lim J, Goh J, Chionh HL, Yap P. Why do patients and their families not use services for dementia? Perspectives from a developed Asian country. Int Psychogeriatr. 2012;24(10):1571–1580. doi: 10.1017/S1041610212000919

18. Wolfs CA, de Vugt ME, Verkaaik M, Verkade PJ, Verhey FR. Empowered or overpowered? Service use, needs, wants and demands in elderly patients with cognitive impairments. Int J Geriatr Psychiatry. 2010;25(10):1006–1012. doi: 10.1002/gps.2451

19. Beeber AS, Thorpe JM, Clipp EC. Community-based service use by elders with dementia and their caregivers: a latent class analysis. Nurs Res. 2008;57(5):312–321. doi: 10.1097/01.NNR.0000313500.07475.eb

20. Janssen N, Handels RL, Koehler S, et al. Combinations of Service Use Types of People With Early Cognitive Disorders. J Am Med Dir Assoc. 2016;17(7):620–625. doi: 10.1016/j.jamda.2016.02.034

21. Draper B, Low LF, Brodaty H. Integrated care for adults with dementia and other cognitive disorders. Int Rev Psychiatry. 2018;30(6):272–291. doi: 10.1080/09540261.2018.1564021

22. Penning MJ, Cloutier DS, Nuernberger K, MacDonald SWS, Taylor D. Long-term care trajectories in Canadian context: patterns and predictors of publicly funded care. J Gerontol B Psychol Sci Soc Sci. 2018;73(6):1077–1087. doi: 10.1093/geronb/gbw104

23. Zhu CW, Sano M, Ferris SH, Whitehouse PJ, Patterson MB, Aisen PS. Health-related resource use and costs in elderly adults with and without mild cognitive impairment. J Am Geriatr Soc. 2013;61(3):396–402. doi: 10.1111/jgs.12132

24. Bugliari D, Carroll J, Hayden O, et al. RAND HRS Longitudinal File 2018 (V2) Documentation: Includes 1992-2018 (Final Release). RAND; 2022.

25. Langa KM, Weir DR, Kabeto M, Sonnega A. Langa-Weir classification of cognitive function (1995 onward). Survey Research Center Institute for Social Research, University of Michigan; 2020.

26. Crimmins EM, Kim JK, Langa KM, Weir DRJJoGSBPS, Sciences S. Assessment of cognition using surveys and neuropsychological assessment: the Health and Retirement Study and the Aging, Demographics, and Memory Study. 2011;66(suppl_1):i162–i171. doi: 10.1093/geronb/gbr048

27. Cloutier DS, Penning MJ, Nuernberger K, Taylor D, MacDonald S, Health. Long-term care service trajectories and their predictors for persons living with dementia: Results from a Canadian study. Journal of Aging and Health. 2019;31(1):139–164. doi: 10.1177/0898264317725618

28. Collins LM, Lanza ST. Latent class and latent transition analysis: With applications in the social, behavioral, and health sciences. New Jersey: John Wiley & Sons; 2010.

29. Sophia R-H, Skrondal A. Multilevel and longitudinal modeling using Stata. STATA press; 2012.

30. Bäck MA, Calltorp J. The Norrtaelje model: a unique model for integrated health and social care in Sweden. International Journal of Integrated Care. 2015;15(September). doi: 10.5334/ijic.2244

